# Facilitators and Barriers to Recruitment and Engagement with a Randomised Controlled Trial in Mental Health Services: experiences of trial participants and intervention providers

**DOI:** 10.1101/2025.10.08.25337563

**Authors:** Georgia Naughton, Brynmor Lloyd-Evans, Jo Billings, Hannah Gray, Gareth Ambler, Nick Barber, Beverley Chipp, Joanne Clare, Maev Conneely, Gamze Evlat, Tumelo Tara Hilton, Manisha Jain, Sonia Johnson, Hameed Khan, Jessica Lashko, Glyn Lewis, Steven Marwaha, Zubair Matin, Ailsa McGuinness, Chandani Nekitsing, Vanessa Pinfold, Prisha Shah, Jerusaa Vasikaran, Martin Webber, Mohammed Zaman, Tanya Mackay

## Abstract

**Background:** Many randomised control trials struggle to recruit and retain participants. Few previous studies explore participants’ experiences of recruitment and engagement in trials in secondary mental health care contexts, and few include the perspectives of control group participants. We addressed this knowledge gap in a sub-study of the Community Navigator trial, a multi-site UK trial of a novel, co-produced, social intervention for people with Treatment Resistant Depression (TRD) in secondary care community mental health teams. We used a mixed methods approach to explore barriers and facilitators to trial recruitment and retention, intervention engagement, and how these factors may persist and inter-relate.

**Methods:** Semi structured interviews were conducted with 17 trial participants and with intervention providers: eight Community Navigators and one supervisor, across four UK sites. An online survey was sent to the first 100 trial participants, which could be completed anonymously. Collaborative framework analysis was used by the research team and lived experience advisory panel (LEAP) to analyse interview, focus group and survey data.

**Results:** Factors helping and hindering trial recruitment and engagement were described in three overarching themes: The Trial Context, Skilled Communication, and The Complexity of Depression and Anxiety. Facilitators included: maximising choice for participants at all stages, a kind and patient approach and striking the correct balance between too much and too little information about the trial. A novel finding was that researchers initiate a “virtuous cycle”, where participants’ initial good experience generates optimism about future involvement in the trial and the experimental treatment. Barriers to recruitment and engagement included referring clinicians providing inaccurate initial information to service users, overwhelming volumes of information, concern about randomisation to the control group, and service users’ internal barriers regarding their mental health.

**Conclusion:** We generated learning for future trials in mental health contexts. Consideration of how to minimise disappointment following allocation to a control group is important. A kind and patient approach from researchers is essential, and a good early experience of the research process can help trial retention and generate therapeutic optimism about the trial intervention.

**Trial registration:** Registered on 8 ^th^ July 2022: ISRCTN 13205972

## Background

Randomised controlled trials (RCTs) are important in establishing if treatments in healthcare are effective, so better understanding of what helps to engage and retain participants in trials is of high interest. Many RCTs struggle to recruit an adequate sample size: research in the UK found that 45% of publicly funded trials required an extension and 80% of industry trials did not meet enrolment deadlines ^1,2^. Challenges regarding participation, retention and obtaining follow-ups are especially common in mental health trials ^3^. Many trials also struggle with an under-representation of participants from minority backgrounds ^4^. Retaining participants is also a common problem for RCTs. These difficulties can lead to reduced statistical power and unrepresentative samples, affecting the reliability, validity and generalisability of the findings of RCTs in healthcare settings.

Recent literature, including two umbrella reviews and a qualitative evidence synthesis, has highlighted a diverse range of factors related to participants’ experiences that influence the success of recruitment in healthcare research ^5,6,7^. The perceived potential for personal benefit, alongside altruism and the desire to contribute to research that may help others in the future, have consistently been identified as facilitators to trial recruitment. Perceived costs of participation – particularly in terms of time and financial burden – along with concerns about risks to health and confidentiality, and an aversion to randomisation have been found to deter enrolment. Additionally, levels of trust in healthcare professionals and researchers, as well as the quality and quantity of information provided about the trial and researchers’ approach to communication are key factors that hinder or facilitate recruitment efforts ^8,9^.

Within the context of mental health trials, there are additional, unique challenges associated with recruiting and engaging participants with mental health issues ^10^. Several existing reviews have explored factors influencing participation among individuals with mental health conditions from the perspectives of both potential participants and referring healthcare professionals ^11, 12, 13^. These reviews have found that many of the general factors affecting recruitment to clinical trials – such as trust and expected costs versus benefits – also apply to populations with mental health issues. However, this population may be particularly deterred from participating in research by the perceived risk of exacerbating mental health symptoms, and the impact of these symptoms on their ability to enrol in and engage with the trial. Stigma associated with the health conditions listed as inclusion criteria can further discourage participation ^11, 13^. Health professionals often share these concerns, which can result in overprotectiveness and gatekeeping behaviours. Participant information sheets and recruitment materials can be inaccessible and off-putting due to the length and complexity required by ethical and research governance ^14^. Moreover, clinicians may lack confidence in introducing research opportunities within clinical consultations, further discouraging them from referring service users into mental health trials ^12, 13.^

Recruitment poses a particular challenge in trials for people with clinical depression ^15, 16, 17, 18^ for many of the reasons listed above. A systematic review conducted by Hughes-Morley and colleagues ^10^ found that challenges included the presentation and impact of depressive symptoms, fear of symptom exacerbation, stigma, and viewing the trial as an inconvenience and burden. A recent feasibility trial by our research group^19^ of a novel, social intervention for people with treatment resistant depression, the Community Navigator programme, reported on specific barriers to engagement and participation with the programme ^20^. It found that severely depressed people often experienced ambivalence and some reservations about signing up to take part in a trial and to engaging with the programme, even when they really wanted the additional support on offer, because it was “emotionally effortful to do so and a major personal challenge” ^20^. Despite the existing evidence base on factors affecting participation in mental health trials, several gaps remain to be addressed. In the most comprehensive review of research in this area to date, included studies were almost exclusively conducted in primary care settings, meaning experiences of individuals with serious mental illness may be underrepresented ^10^. Additionally, previous research has focused primarily on how to increase recruitment numbers, rather than on the quality of the recruitment process experienced by participants, and has often overlooked retention in mental health trials ^10, 13^. The methods used to study factors affecting recruitment have been fairly homogenous, generally involving qualitative interviews or focus groups. In a review of factors affecting recruitment to clinical trials, Houghton and colleagues ^6^ observe that there has been little to no mention of Public and Patient Involvement (PPI) in studies published so far.

The current, multi-site randomised controlled trial of the Community Navigator programme^21^ provided an opportunity to address some of these gaps in knowledge and further explore barriers and facilitators to trial recruitment and retention, intervention engagement, and how these may persist and inter-relate. The Community Navigator trial is a multi-site UK National Health Service (NHS) trial of a novel, co-produced, complex intervention for people with Treatment Resistant Depression (TRD) in secondary care community mental health teams. The intervention is designed to reduce symptoms of depression by helping people develop meaningful social connections and thus reducing loneliness (Stefanidou et al., 2023). The intervention and trial process are summarised in Figure 1.

**Figure 1.**
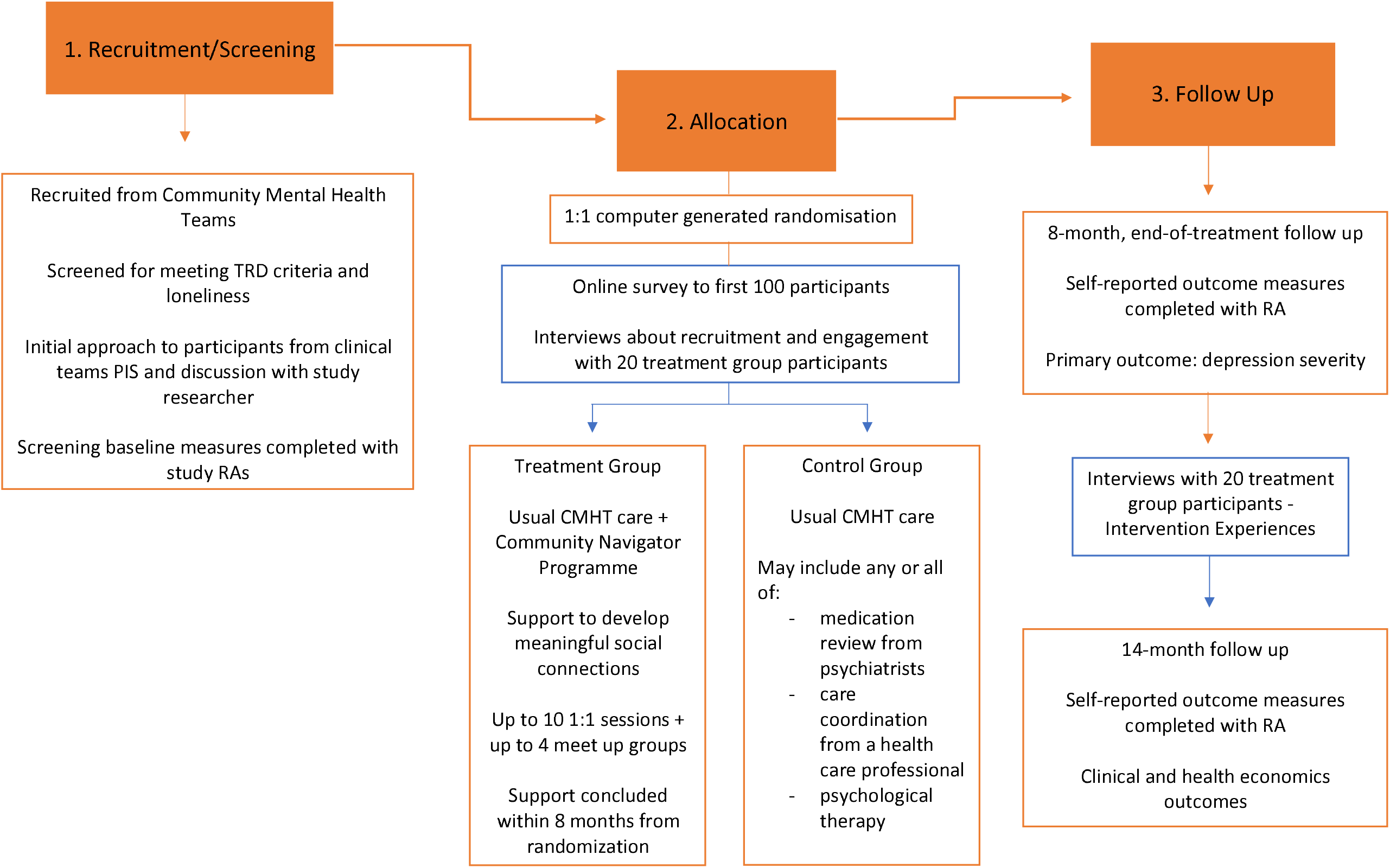
Within the Community Navigator programme trial, we used qualitative methods to explore participants’ experiences of recruitment to the trial and initial engagement with the programme, both to identify any rectifiable barriers, and to provide transferrable knowledge about how to support recruitment and engagement of people with mental health problems, specifically TRD, in future mental health trials.

## Methods

This was a multi-method qualitative study using data obtained through interviews, focus groups and an online survey. Our research involved three methods of data collection:

1. Interviews with intervention group participants
2. Interviews or focus groups with Community Navigators (the trial intervention providers) and their supervisors.
3. An invitation to the first 100 trial participants (intervention and control group) to complete an anonymous online survey, providing feedback on the recruitment process and experience of engaging with the intervention (where applicable) through structured and free text questions.

### Setting

We recruited participants from Community Mental Health Teams or equivalent secondary mental health services in four NHS Trusts across England which were taking part in the Community Navigators trial. Two were based in London, one in the Midlands and one in Northeast England. The four sites included areas with high ethnic diversity, urban and semirural areas, and both affluent and deprived areas. (Two further trial sites were added subsequently, after the end of the data collection period for this qualitative study.)

### Participants

For the qualitative interviews, we sought to recruit 20 service user participants who were allocated to the intervention arm of the trial (5 from each site). Our sampling followed purposive principles: we aimed to recruit participants from all the initial four participating NHS Trusts, to include demographic diversity in our sample (gender, age and ethnicity) and to prioritise participants who had disengaged from the programme (who had been allocated to the intervention group, but for various reasons did not continue seeing the community navigator). We sought to recruit participants a few weeks after enrolment into the study, when experiences of the recruitment process and initial engagement with their Community Navigator were still recent. All engaged and disengaged participants had previously given informed consent to being approached by the study researcher for the qualitative interviews.

We also sought to conduct separate interviews or focus groups with all the Community Navigators (maximum three per site), and their supervisors, from each of the four sites.

An online survey was distributed via email to the first one hundred service user participants recruited and randomised into the trial, from both intervention and control arms. Participants completed the online survey anonymously.

### Materials

#### Interviews

Separate topic guides for each participant group (see Supplementary File 1) were collaboratively developed by the lived experience advisory panel (LEAP) and a peer researcher – an employed researcher who uses their own lived experience of mental health difficulties to inform their work ^22^ - in conjunction with the wider research team. The guides were used to guide the semi-structured interviews with service users in the intervention group, Community Navigators and supervisors. The topic guides explored participants’ experiences and expectations of the programme from the recruitment process, obstacles to initial participation and any barriers to subsequent engagement.

#### Survey

A digital online qualitative survey (see Supplementary File 2) containing structured and free-text questions was collaboratively developed by the peer researcher and LEAP, with the wider research team, to explore service user participants’ views on the recruitment process and materials. The survey was designed using the Qualtrics software programme with branching questions, so that respondents who reported being allocated to the intervention group were also asked about barriers to engagement with the intervention. Participants from the control group were also asked about their feelings towards being allocated to the control group.

### Procedures

#### Interviews

Service user participants in the intervention group, Community Navigators and supervisors were invited to participate in a qualitative interview or focus group by the peer researcher. The researcher identified themselves to participants as a peer researcher with lived experience of mental health problems. Prospective participants were provided with separate information sheets and written or audio-recorded verbal consent to participate was obtained prior to interview. The participants were able to choose whether they would like the interview to take place face-to-face, online via video call or over the phone. Service user participants were reimbursed for their participation with a £20 gift voucher. Interviews and focus groups were audio-recorded and transcribed verbatim by a GDPR-compliant external professional transcription company. All identifying features of participants and services have been removed to ensure anonymity. Anonymous participant identifiers are used in the results.

#### Survey

The qualitative survey link was distributed via email the first 100 service user participants recruited onto the trial. An information sheet was included at the start of the survey, accessed through the online link. Respondents consented to take part by clicking a consent button at the end of the information sheet and proceeding to the survey questions. No financial reimbursement was offered to survey respondents.

### Analysis

The interview and focus group data were firstly used to provide rapid feedback to the study team to make any adjustments to the recruitment process and programme as necessary. Full analysis followed.

#### Interviews

After all interviews and focus groups were completed and transcribed, a two-stage collaborative framework analysis was used to analyse the relevant data ^23, 24^. This followed a process which was also previously used in the Community Navigator feasibility study ^20^. The research team involved in the collaborative analysis included two lived experience researchers, five lived experience advisory group (LEAP) members with personal experience of anxiety, depression, and loneliness, four senior academic researchers with clinical qualifications in social work and psychology, four research assistants, one MSc student and one experienced Community Navigator involved as a co-applicant in the current study.

#### Survey

Responses to the structured survey questions were descriptively summarised once the survey was closed to respondents. Results from the free text questions were incorporated into the same process of analysis alongside the qualitative data from interviews and focus groups. Responses to closed questions from the survey are reported in supplementary file 3. In the first stage of our analysis, a hybrid (in-person and videoconference) collaborative analysis workshop was held where the research group worked in teams to selectively extract and index data from a sample of transcripts. The teams explored, compared and then discussed issues relating to experiences of recruitment and initial programme engagement, as well as potential barriers, identifying potential categories for the analytic framework. The peer researcher and members of the research team subsequently worked on extracting and indexing data for the full set of transcripts. All extracted data was then entered into the framework using Microsoft Excel. Separate frameworks were created for service user, Community Navigator, and supervisor data, using the collaboratively developed codes.

At the second stage of analysis, the peer researcher reviewed the data within the framework analysis and synthesised the key themes. Provisional themes from each of the frameworks were entered into a theme map and table formats which illustrated how related codes and data interlinked. These were shared with the wider research team for review with reference to one or more anonymised interview transcripts. Feedback was also provided regarding whether the refined set of themes and codes encompassed all the relevant data, which was used by the peer researcher to further refine the results (see Supplementary File 4).

## Results

### Qualitative Interviews

Semi-structured interviews were conducted with seventeen service user participants; including two who had formally withdrawn from the intervention and fifteen who were still taking part in the intervention at the time of their interview.

Interviews and focus groups were also conducted with eight Community Navigators and one supervisor across the four different sites. These included four Community Navigator individual interviews, two focus groups with Community Navigators, each with two members, and one supervisor interview. Participation in interviews or focus groups depended on the availability of the Community Navigator: not all Community Navigators were able to make the scheduled focus groups and chose to do an individual interview at a later date. All interviews and focus groups took place between December 2022 and February 2024.

The characteristics of the service user and Community Navigator participants involved in the qualitative interviews are described in Table 1.

**Table 1.** Socio-demographic Characteristics of interviewees.

| **Participants** | **Gender** | **Age**<br>**(mean,**<br>**s.d.)** | **Ethnicity** | **NHS Trust** |
| --- | --- | --- | --- | --- |
| **Service user (n=17)** | Male: 8<br>(47%)<br>Female: 8<br>(47%)<br>Non-binary:<br>1 | 42.4 (11) | White British: 12<br>(71%)<br>Mixed/ Multiple<br>ethnic groups: 2<br>Asian/ Asian<br>British: 2<br>Other ethnic<br>group: 1 | London A: 3<br>London B: 2<br>Midlands: 4<br>Northeast: 8 |
| **Community Navigators**<br>**& Supervisors N=7**<br>**(two further**<br>**participants did not**<br>**provide demographic**<br>**data).** | Female: 7<br>(100%) | 44 (11) | White British: 5<br>White other: 1<br>Asian/Asian<br>British:1 | London A: 1<br>London B: 2<br>Midlands: 1<br>Northeast: 3 |

**Table 2.** Themes and subthemes derived from qualitative interviews & survey.

| Theme | Subtheme |
| --- | --- |
| The Trial Context | Initial Information |
|  | Randomisation |
|  | Organisational challenges |
| Skilled Communication | Kindness and patience |
|  | Flexibility and choice |
| The Complexity of Depression and Anxiety | Internal barriers |
|  | Disengagement |

### Qualitative survey

The qualitative survey was sent out via email to the first one-hundred participants recruited to the trial, between September 2022 and May 2023. Twenty-three participants completed the anonymous survey, thirteen in the intervention group and ten in the control group. We did not collect participant demographics during this survey due to its anonymous nature.

The survey responses, and feedback from interview participants, prompted some adjustments to the researchers’ training and how the trial was explained to participants. These related to reinforcing and double-checking participants’ understanding of the implications of being allocated to the control group; providing continuity in who contacts participants and discusses participating in the trial wherever possible; and further communication with clinical teams to ensure consistent messaging about the nature and limits of the support offered through the trial intervention. Further information is provided in Supplementary File 5.

Themes from the initial stages of analysis were combined and distilled into three overarching themes: (1) *The Trial Context* (2) *Skilled Communication* (3) *The Complexity of Depression and Anxiety*. These themes and the subthemes within them are summarised in table 3.

### The Trial Context

Recruiting and engaging service users in a trial focusing on treatment resistant depression and anxiety involved several challenges. Service user trial participants reported being provided with accurate information about the trial, time constraints, persisting mental health symptoms and randomisation into the control group. However, support from the research assistants facilitated recruitment.

#### Initial Information

Service users were first approached about the Community Navigator programme by a member of the clinical team, usually one of their treating clinicians, a research assistant working in their NHS trust or the Community Navigator themselves. The importance of being provided with accurate initial information about the trial from their clinician was emphasised by service user participants, enabling them to understand exactly what the research would involve.

> *“Yes, they explained it, everything about it. Yes, I wasn’t in doubt of anything. It was really, really in depth. Yes, I knew exactly what was happening*.*”* (SU10).

The study researchers provided the participants with a lot of information when they were first recruited onto the trial, with the awareness that this could be quite overwhelming for people who were experiencing anxiety and depression. Some participants described a positive recruitment experience where they were provided with the right balance of enough information to consent, without feeling overwhelmed. The majority of the participants who completed the survey, also stated that they found the amount of information provided during the recruitment stage to be “the right amount”.

> *“No, I feel like I had enough information*…*but sometimes I can get overwhelmed with too much information about it, I was given enough information that I understood what it was, but didn’t need to know every single intricate detail. But also knew that I could ask questions if I needed to*.*”* (SU5)

However, one participant described having a negative experience during the early stages of recruitment due to feeling overwhelmed by the amount of communication and information provided by the study researchers, highlighting a sensitive balance between providing sufficient information without it becoming overwhelming for the participants.

> *“There was a lot of communication at first. It was quite, if I’m honest, bombarding. And I hadn’t seen anybody at that point, I was like, “Wow*.*” I found it a bit overwhelming. Because I didn’t really know*… *When I was getting people calling me, I wasn’t aware, at the time, these were the people*… *I didn’t know I was going to get called up all the time like that, so I found it quite overwhelming, because I really don’t like the telephone”* (SU8)

One participant shared that they felt the information provided by their clinician was inaccurate regarding the Community Navigator’s role. The participant’s initial expectations for more holistic support endured throughout the recruitment process and in turn caused them distress, due to the support provided by the intervention focusing solely on support with social connections and thus not meeting their expectations.

> *“My expectation was that maybe they’re [Community Navigator] like a support worker*, … *dealing with my needs every day*…*And [the clinician] didn’t care, didn’t listen to me, and didn’t help me. They [the Community Navigator] kept saying, “Now, you have a care coordinator. Ask her [Care Coordinator]*.*” And that’s why it made me more mad, it made me more sad. And I just came out of the research, I said, “I want to withdraw”, because it’s going to be more complicated for me*.*”* (SU16)

The survey responses suggested that the majority of the participants found the written information provided during the recruitment stage to be clear and understandable. However, two participants who were experiencing difficulties with concentration and memory reported finding it hard to take in all the information provided during the early recruitment stages of the trial. They found it essential to have the research assistant present when they were reading through the participant information sheets to help explain anything they were struggling to understand. A requirement for some for large amounts of detailed written information was also having someone to speak to and ask questions about the written information.

> *“Yes, it was easy to understand, but it is just my concentration isn’t very good, so it took me a little while, that is all*.*”* (SU15)
>
> *“A bit overpowering, because in your head, sometimes it gets a bit foggy, you know?*… *just make sure that there’s someone there that can explain it properly and help fill the paperwork in, that’s the only thing*.*”* (SU4)

#### Randomisation

Responses to our online survey indicated that some participants who were randomly assigned to the control group reported being distressed because of their allocation. For example, one participant expressed feelings of disappointment and rejection, which appeared to compound previous experiences of the mental health system.

> *“Disappointment after rejection time after time have negatively impacted my mental health leading to self-harm AGAIN and more time in A&E*.*”* (Survey response 6)

Another service user participant expressed concern about the impact of their disappointment in relation to being allocated to the control group on their current mental health. Even though participants were allocated at random, it can still feel personal when not chosen for the intervention.

> *“Was worried about the negative impact on me and disappointment I would feel if not chosen*.*”* (Survey response 6)

#### Organisational Challenges

Community Navigators were newly employed for the research study. There were initial logistical challenges that led to issues engaging with the participants. These included access to work equipment, such as a mobile phone.

> *“I think one of the things if I could just say, was just me not having a work phone for a while. That was a challenge because sometimes they couldn’t really get to me or if I called them, they couldn’t call me back because I would call from a private number. So they couldn’t get back to me and confirm appointments or, sort of, have that contact with me, or receive text message reminders*.*”* (CN2 / Focus Group 1)

The part-time nature of the Community Navigators’ employment to deliver the trial intervention limited how flexible they could be, which was negatively experienced by some participants.

> *“*…*because she works part -time, obviously, she’s not contactable, what is it? Thursdays and Fridays? And whilst I’ve been okay to meet, I have to reschedule. And, yes. I mean, it is a bit of a pain having to make a mental note set a reminder to do it Monday. But I suppose you can’t really do much about that*.*”* (SU11).

Part-time working also made it more difficult for Community Navigators to communicate with their colleagues and other members of the clinical team, to gather required information before meeting participants.

> *“I’m in the same office as one person but working on a different day and I know they work the same day but in different offices*… *“for example, this week, because I’m collaborating with my other two Community Navigators, and seeing my person on Friday, it means I’m working three days*.*”* (CN1/Focus Group 1)

### Skilled Communication

The participants and spoke about how compassionate communication from both researchers and intervention providers was key to engagement in the trial and the programme. These positive interactions involved kindness and patience, balance, and choice.

#### Kindness and Patience

Being treated with kindness and patience from the first interaction with the research team created a positive initial impression of the programme. Having a positive experience when interacting with the research team set the tone for later interactions and created a virtuous cycle for the service users, meaning participants expected further communication with other staff in the trial to be of a similar positive nature.

> *“That experience for me was good, it was very good because I was always treated with patience, with kindness. Yes, that was important to me after being so hesitant because of the reasons that I said before. I took the decision, I get the first step, because she [the study researcher] explained me clearly. She was very kind. She was patient. So, for me it was a good experience”* (SU3)

Positive interactions with the research team led service users to believe that they would be treated in such way throughout the programme and created more optimism about engaging with the intervention.

> *“I usually normally feel nervous, but I have to be honest, I was expecting my Community Navigator will be understanding and, yes, a nice person. And when I met her, (Community Navigator 3) it was, yes, it’s been okay, she’s a really nice person*.*”* (SU3)

The Community Navigators were described as “understanding” after their initial interactions. Participants explained that they felt more listened to by the Community Navigators than they were sometimes used to during appointments for their mental health care.

> *“I think the Community Navigator that I’m with was very understanding. It was, so you feel like you are being listened to really, so that is good*…*Yes. I thought it was probably going to be one of those on treatment team visit where they just ask you routine questioning*.*”* (SU15)

#### Flexibility and Choice

The study researchers provided the participants with a choice in format of how they would like to receive the participant information during the initial stages of recruitment, which helped them to make an informed choice to whether they would like to take part in the trial.

> *“Yes, the information was presented verbally on the phone, and then on paper form. I had everything I needed to make an informed choice. No, I would say, yes, wouldn’t really say there is anything else you could do differently or better*.*” (SU10)*

The participants valued the researchers giving them choice regarding the pace at which they signed up to the study and completed the onboarding process.

> *“She was lovely, really friendly, asked if I had any questions all the way through, didn’t feel rushed at all into making a decision there and then*.*” (SU7)*

They also valued having a level of choice in the pace at which they moved through the trial, without being pushed in any direction too quickly and feeling like they had to meet certain goals and timelines.

> *“She said that she’s there for me. She doesn’t want to push anything on me. It’s more about just guiding me, rather than pushing anything on me. So, yes, she’s just-yes, just very understanding*…*”*(SU13)

This element of choice also applied to ways in which the participants could communicate with their Community Navigator. Having this flexibility around communication helped participants to stay engaged in the intervention.

> *“And the facility that I can text her, or for example if I miss a call, I can text her or she can reply to me, she can phone me back, something like that. Simple things like that, so it’s been okay*.*”* (SU4)

### The Complexity of Depression and Anxiety

Symptoms associated with depression experienced by the service users were found to sometimes create barriers to their initial involvement and recruitment into the study. Service users with comorbidities and experiencing other socioeconomic issues could also make it difficult for the Community Navigators to know where the boundaries were between their role and supporting them further with for example housing or employment advice.

#### Internal barriers

Service users reflected that their current mental health issues can sometimes make it difficult to fully take in all the information they were provided with by the research assistant and Community Navigator. Service users reported that they sometimes struggled to feel motivated to be able to fully engage with the programme.

> *“Because my mental health isn’t there basically, just isn’t there on point, and then I end up not engaging as much as I probably should, or I wasn’t able to respond and stuff like that*.*”* (SU15)
>
> *“Sometimes when you have a depression, high level of anxiety and stress, you can’t focus on what you’re hearing, what you’re learning. Sometimes I need to ask the people to repeat the words many times*.*”* (SU16)

When participants were asked as part of the survey to whether they had any concerns about taking part in the trial, some voiced concerns around how their anxiety may act as a barrier to them taking part in the trial.

> *“Just my anxiety about going out”* (Survey response 13)
>
> *“That my anxiety would prevent me and my negativity would talk myself out of it”* (Survey response 19)

Timing was an important aspect, regarding whether the service user was at a time in their life where they felt able to take on the commitment needed to engage in the trial and whether making connections was a current priority for their wellbeing.

> *“But, you know, it’s very hard, when you’re depressed, or you have a mental health problem, to go and find your hobbies, or anything. And that’s why it was really hard for me to continue with the care navigator. Just thinking about it, what I am going-because at that moment, I didn’t enjoy anything, you know? And there is no point for me to talk about socialising”* (SU16)

Negative self-beliefs and low self-esteem which often coincide with depression were also expressed. For example, one participant talked about feeling like they didn’t deserve to take part in the study.

> *“I think my only thing was, I guess in some ways it’s probably just my own brain but, “Am I too well for this study”, kind of stuff”* (SU5)

#### Disengagement

Community Navigators reported challenges in relation to organising consistent appointments for their sessions and trying to engage the people they were supporting in the programme.

> *“But every time, she would cancel it due to physical health problems or physical health appointments. When she did withdraw from the project, I did wonder if there was anything I could have done more at that point to, kind of, try to get her to meet in person. And there was part of me that wondered, if we had been able to meet in person, if that might have helped her to stay in the trial*.*”* (CN7)

They explained that participants not being able to commit to their appointments made it difficult to create a work schedule, even when they were trying to be as flexible as possible.

> *“I guess also that can be quite hard because I think sometimes I found that part also-because they keep postponing, I can’t make a plan. So I don’t know what to do with my schedule sometimes. I understand that it is important to be flexible and it is part of our role, but then sometimes that has complicated my life, I guess*.*”* (CN8)

## Discussion

### Main Findings

Our qualitative study identified several facilitators of recruitment and engagement in this RCT for people with severe depression and anxiety. These included allowing the service users to have choices wherever possible, treating them with kindness and patience throughout, and striking the correct balance between providing too much and too little information regarding the trial. Materials written in plain, accessible language accompanied by explanations from the researcher were also important. Our study has also highlighted many barriers to recruitment and sustained engagement, such as referring clinicians providing inaccurate initial information to service users leading to unmet expectations, researchers providing overwhelming volumes of information, concern about being randomised to the control group, organisational and technical challenges such as navigator phones not working and trial participants’ internal struggles regarding their mental health.

Our results broadly fit within the existing literature such as it being key for researchers to have ample time to allow for participants to build trust, allow for asking questions and getting clarification on the large amount of written text. Researcher’s approaches were key to reducing attrition and improving experiences. A similar finding was also made by Azad and colleagues ^25^ who also mentioned the importance of flexibility and the human encounter being critical. Our findings also suggest that the quality, quantity and accessibility of the information provided about the trial and the researcher team’s approach to communication were found to be essential in hindering or facilitating recruitment, which was also highlighted in recent reviews investigating the successes of recruitment in healthcare ^5, 6, 7^. These reviews also identified barriers similar to those found in the current study, including concerns associated with risks to health. Furthermore, our findings relating to participant difficulties in reading and digesting the trial information sheets and staying engaged in the intervention, supports that of reviews exploring factors influencing recruitment and engagement in trials specifically in the context of mental health trials. These reviews highlighted that people with mental health issues may be particularly deterred from participating and engaging in research trials due to their symptoms, and being concerned that their symptoms would prevent them having an impact on their ability to enrol and engage with trials ^10, 11, 12,13^. The fear of being placed in the control group was also a reason why some people declined to take part in the first place, similarly to findings by Fallowfield and colleagues ^26^.

Service user participants reported that their initial conversations with the research assistant during the onboarding process were positive as the research assistants were warm and considerate. A novel finding in our study related to these initial positive experiences creating a ‘positive contagion’, allowing them to expect that future researchers and the Community Navigators would also treat them well, therefore these positive experiences motivated the service user to stay engaged with the trial. Participants experienced this positive communication throughout the trial, allowing them to feel comfortable around their Community Navigator which enabled them to share their thoughts and feelings with them and engage in the intervention.

As our survey was also completed by control group participants, we were able to capture their feelings towards not receiving the intervention. Investigating the views of the control group is not a common occurrence in trials, however, by doing this we were able to find that a number of participants were upset by their allocation and displayed varying degrees of distress in their responses. This echoes findings in existing reviews which highlight people often not wanting to participate in trials due to the element of randomisation and the chance of not receiving the intervention ^5, 6, 7^. Our study suggests that people who are already depressed, may find allocation to a trial control group particularly challenging, experiencing a loss of hope and frustration due to not receiving a potential helpful intervention.

### Strengths and Limitation

This study triangulates information from several stakeholder groups and information sources to explore influences on trial participation and engagement in a mental health intervention, including participants allocated to treatment and control groups. Interviews and focus groups provided rich contextual information about what helps and hinders trial recruitment and engagement with an intervention. Our anonymous, online survey may have elicited views from some trial participants who would not be willing to take part in an interview, as well as participants in the control arm of the study. Studies do not routinely ask participants to provide feedback on their recruitment experiences, but doing so was consistent with our co-production ethos and added considerable value to the study. However, a limitation to this was the small sample size and self-selecting nature of the recruitment, which may not represent the breadth of experiences. The study ensured that feedback from the interviews and survey informed recruitment processes in our trial, which may have helped recruitment. Our collaborative approach to analysis, involving people with mental health experience and practice experience, enhanced the robustness of our analysis approach and resulting findings.

Our collaborative approach to data analysis and theme development, including multiple perspectives, helped avoid undue subjectivity in developing the findings from this study. The research team included two lived experience researchers, five LEAP members, with personal experience of living with anxiety, depression, and loneliness, four senior academic researchers with backgrounds in social work, mental health, and psychology, four research assistants, one MSc student and one Community Navigator from the feasibility trial. As a peer researcher with lived experience similar to that of the service user participants, GN was able to use an empathetic approach towards communication with the participants during recruitment and during the interviews. Having this level of empathy allowed GN to be understanding of any concerns and issues they experienced during the intervention. GN’s lens also influenced how the data was viewed during the analysis, allowing a deep understanding of the participants experience due to its relatability. Some of the LEAP reflected that they have similar lived experiences to the service user participants in the study, including experiences of loneliness, anxiety, depression, neurodiversity, OCD, social withdrawal and using community mental health services. They also have experience of being involved in peer support projects and other research projects as facilitators and recipients. They have interests in how mental health issues and trauma can affect our ability to connect with people and create barriers to being part of communities. They reflected that their positionality meant they could empathically put themselves into the shoes of the participants throughout the stages of the trial. Having this understanding of social withdrawal allowed them to inform the research by making suggestions for improvements to the Community Navigator training and in turn bettering the experience for the service users, for example helping to meet different communication needs and ensuring researchers’ roles were explained very clearly. This lens also impacted the creation of the interview schedule and the analysis of the interview transcripts, by providing insight on what are the most crucial elements of the process of participating and engaging with the trial and its intervention for the service users.

We over-recruited from the North-East England study site, reflecting their high levels of participant recruitment to the trial itself. The demographic characteristics of our interview participants broadly reflect those of the trial participants overall and the services from which we recruited. However, we were unable to sample for maximal variation on demographic characteristics and included relatively small numbers of participants from minority ethnic groups, for whom barriers to participation in research can be particularly marked ^13^. To ensure participants’ confidence in the anonymity of our online survey, we did not collect any demographic information, so we cannot tell how far survey respondents’ characteristics reflect the overall trial population. This limitation means are our results do not provide insight into bespoke approaches to recruitment that might increase participation specific demographic groups.

Due to time constraints, we recruited 17 trial participants for interviews, rather than 20 as planned. We had initially intended to prioritise participants who withdrew from the trial or disengaged from the programme. However, withdrawal rates in the trial were low, and only 2 of 14 people who withdrew during our qualitative data collection period agreed to be interviewed. We therefore had limited information to explore participants’ reasons for withdrawal.

It may be beneficial for future trials to explore recruitment and engagement experiences as findings from our trial may not be typical, such as the low response rate to the online survey which in the context of our study could be due to internet access, differing levels of digital skills, sub-optimal user-interface design, participants’ mental health symptoms i.e. tiredness, lack of motivation, or lack of rapport with the research who contacted them with the survey. A financial incentivisation could also be considered for this online survey element of the study. It may also be beneficial to interview clinicians on their experiences of referring patients into trials, how it is done and how it is framed, and the reflections of the research assistants who recruited participants, to investigate any facilitators and barriers at this stage too.

### Implications

Four recommendations from our research for future trials regarding the trial recruitment and engagement process are:

### Researcher Approaches

Researchers need to be friendly, patient, flexible, trauma informed and encouraging within the limits of what is reasonable within the time frame and requirements of the trial, to optimise participants’ experience of the whole recruitment process ^27^. This will not only make people feel inclined to participate in this and future research but will also set the trial intervention up on a good footing, with people feeling optimistic about a good experience. Recruitment is a critical first step in building research teams that can actively engage in the approaches and ways of working detailed in this paper. Further, the importance of his way of working needs to be built into the researchers’ training and workload, so they have the resources and time to do this well. Required Good Clinical Practice training ^28^ currently focuses on processes and data security, rather than skilled and supportive communication. In our trial, the training for researchers also included role playing recruitment with LEAP members and receiving feedback, which we recommend. Study leads should ensure that anxieties around recruitment targets do not lead researchers to pushing service users or not giving them inadequate time and space to make an informed choice, which could lead to higher subsequent dropout rates due to feeling they have been persuaded to participate rather than being personally motivated. Creating a virtuous circle by providing positive initial experiences with the researchers during recruitment gives a good foundation and anticipation of a positive experience in the intervention

### Making Information Accessible Throughout

Information needs to be clear, in plain language free of jargon or unusual words, accessible, and reiterated consistently. Lived experience involvement in developing the recruitment materials is key to achieving the right balance of information without overwhelming the service users. There needs to be sufficient time, on more than one occasion where needed, for the researchers to go through and discuss the information with services users too. It is not enough to rely on written information alone, or a one-off explanation, therefore good clear information should be provided to clinical teams, who may be making the first approach. A number of our interview participants mentioned finding it challenging to digest and retain the relatively complicated information required to explain a randomised controlled trial. Research assistants therefore need good training in assessing mental capacity to make an informed choice about trial participation, as our findings suggest people with depression can find it challenging to engage with and retain information. This should include proactively checking participants’ understanding of trial information and offering multiple opportunities to discuss and clarify information, which can ensure people are able to provide informed consent to enter a trial and was valued by our participants. These adjustments could also be usefully considered with neurodivergent populations and other mental health populations who may experience cognitive issues, such as psychosis and dementia.

### Clarity from Clinical Teams

In our trial there were some inconsistencies in how the trial was presented to service users by the various people involved in the recruitment stages. This taught us that researchers need ongoing attention and engagement with clinical staff make sure that clinical teams have a clear understanding of what the intervention is and is not ^29^. However, this may also reflect the current pressures on mental health services, as clinicians may feel pressure to access any additional support to help address a range of unmet needs for their clients and take the pressures off usual care. This suggests a need to carefully monitor what usual care consists of for both services users in the intervention and control group and to try and check if getting allocated to the intervention means a reduction in their usual care, which isn’t what they signed up for and could undermine the trial. It may therefore be useful to hear the views of clinical staff involved in making the first approach to potential participants in further studies, to understand their experiences and how a trial team can best support them.

### Efforts to Mitigate Control Group Allocation Disappointment

Our qualitative study also has implications for the design of future trials regarding the nature of the control condition. Our study suggests that allocation to a treatment as usual control group may be upsetting for some participants, at least among participants who are severely depressed ^30^. However, it would be beneficial for further research to investigate the mitigation of impacts of allocation to a treatment as usual control condition in trials of psychosocial interventions where participant-blinding is not possible, especially within depression and anxiety populations, who may be sensitive to perceived rejection and disappointment. In our trial we attempted to mitigate this by clearly stating in written and verbal information that they might not receive the intervention when taking part in the trial. Research assistants should also ask the service users if being allocated to the control would be disappointing for them and if so, would the disappointment be manageable, to avoid causing distress. This can become a standard part of helping the person to weigh up the pros and cons of taking part in the trial. The research assistants also ensured that they did not over sell the intervention element and didn’t mention the financial support with social connections which Community Navigators could access for people in the intervention group. They also emphasised equipoise: that we don’t know that the novel treatment is helpful. Despite inevitable eagerness to recruit participants, researchers reiterated the chances of being allocated to the control group, so it wasn’t a shock. They emphasised to participants the support on offer to the control group (receipt of e a booklet with many listed community activities, and the continuation of usual care). We arranged phone calls with all participants to inform them of their allocation status, rather than doing this via email or text, so a member of the research team would be there to support them if they were disappointed with their allocation.

## Conclusion

RCTs continue to hold a critical role in evaluating new interventions in mental health care. For high quality, pragmatic trials, robust and diverse study recruitment is essential. However, recruitment is complex, and often context dependant. Our study suggests that recruitment and engagement to RCTs involving people with depression and anxiety can be challenging due to internal barriers and participants feeling overwhelmed with essential trial information. However, our findings also highlight that there are ways in which the research team can help to reduce these challenges and improve recruitment and retention, such as treating participants with compassion and patience throughout and providing flexibility and choice to improve recruitment and engagement. These early experiences can set the tone for participants, creating virtuous circles of positive expectations and experiences throughout the trial process.

### Additional Files

Additional File 1: Interview topic guides

Additional File 2: Survey questions

Additional File 3: Survey responses to closed questions

Additional File 4: Thematic Framework

Additional File 5: Feedback adjustments

## Data Availability

The datasets generated and/or analysed during the current study are not publicly available in line with the information provided to participants, but deidentified data are available from the corresponding author on reasonable request.

## Abbreviations

GDPR: General Data Protection Regulation
LEAP: Lived Experience Advisory Group
PPI: Patient and Public Involvement
RCT: Randomised Controlled Trial
REC: Research Ethics Committee
TRD: Treatment Resistant Depression

## Declarations

### Ethics approval and consent to participate

The trial, and the accompanying qualitative investigation reported in this paper, were approved by the South Central Oxford B Research Ethics Committee on 30th March 2022 (REC reference 22/SC/0064). Written, informed consent to participate in qualitative interviews or focus groups was obtained from all service user, Community Navigator and supervisor participants. Participants enrolled in the trial consented to take part anonymously in the online survey by following a link from an invitation email, reading an online information sheet, and clicking to consent and continue to complete the survey.

### Consent for publication

All participants have consented to publishing results, both on a group level and individually, given adequate anonymisation. All data has been analysed with respect for confidentiality. This information is included in the Participant Information Sheets and consent has been given accordingly.

### Authors’ Contributions

GN is the peer researcher on the Community Navigator trial who conducted the qualitative interviews. BLE, the Chief Investigator and SJ, the co-Chief Investigator conceived the study. GA, JB, SJ, GL, BLE, TM, SM, ZM, VP, and MW are the study applicants: they developed the proposal and trial design. GN led the qualitative framework analysis, with support from BLE, JB, NB, BC, MC, GE, TTH, HK, ZM, CN, VP, PS, JV, MW, MZ and TM. GN drafted the manuscript with help from BLE, JB, TM, MC and HG. All authors reviewed and approved the final manuscript.

## Acknowledgements

Thank you to all the participants, lived experience advisory panel members, involved clinical services and NHS Trusts’ Research Departments for their contributions to and support for this trial.

