## Supplementary Files 1-5 for "Facilitators and Barriers to Recruitment and Engagement with a Randomised Controlled Trial in Mental Health Services: experiences of trial participants and intervention providers"

1. **Interview topic guides**

### Internal Pilot Qualitative Interviews

### Trial participants

**This is a flexible guide to help build a conversation. Not all questions will need to be asked and may not be relevant to all interviewees. Interviewers will paraphrase question to best suit the person they are speaking with.**

**The purpose of this guide to is to *explore people’s experiences of the programme from the recruitment process, obstacles to initial participation, and any barriers to subsequent engagement.***

1) Can you tell me how you found out about the Community Navigator programme?

Probe:

Who introduced the study to you? Explore relationship with them and possible influence on how person felt about the project.

2) How was the Community Navigator programme explained to you?
3) How well did this person introduce the study to you?

Probe:

Did you feel they gave you enough/to much or to little information?

Were you able to ask all the questions you wanted to make a decision?

4) Was there anything unclear or off-putting during the recruitment? Can you tell me about this?
5) Is there anything you wished you had known before you signed up?
6) We had lots of information to give you – flyers, information sheets, consent forms to sign. Can you tell us about how easy or difficult it was to read and complete these?
7) Looking at the recruitment process you went through, how do you think we could improve our approach so that other people will also take part?
8) How did you find your first meeting with the research assistant to complete the initial assessments?
9) What interested you most about the Community Navigators programme?
10) How long did it take you to decide to sign up for Community Navigators? What were the things you considered in making this decision?
11) Did you have any concerns about starting the Community Navigator programme?
12) Can you tell me what your expectations of the Community Navigator programme were when you signed up?
13) Were these expectations been met so far? Why/why not?

Probe: Has anything come up/happened that you weren’t expecting, good or bad?

14) Can you tell me about how you found the process of being connected with a Community Navigator?
15) Did you meet or speak with your Community Navigator? - Can you tell me why not if you haven’t?

**If they haven’t met their Community Navigator:**

What could be changed to help you connect with the Community Navigator?

**If they have met their Community Navigator:**

Can you tell me about how you have communicated so far? (for example have you spoken via telephone or videocall?) How has this been for you?

Have you encountered anything that has stopped you from engaging with your Community Navigator? If so, can you tell me about this?

16) Thinking about your experience so far on the project, can you tell me about anything you think we could improve on?

### Internal Pilot Qualitative Interviews

### Community Navigators

**This is a flexible guide to help build a conversation. Not all questions will need to be asked and may not be relevant to all interviewees. Interviewers will paraphrase question to best suit the person they are speaking with.**

**The purpose of this guide to is to *explore people’s experiences of the programme from the recruitment process, obstacles to initial participation, and any barriers to subsequent engagement.***

1) Can you tell me a bit about what interested you about becoming a Community Navigator?
2) How did you find the training?

Probe:

Do you think we could improve the training in anyway?

3) How did you feel after training, about taking on this role and seeing clients?

Probe: confident? prepared?

4) Do you feel the Community Navigators programme is going to help clients address loneliness and depression? Why or why not?
5) Do you have any concerns about how the programme is going to work in general?
6) How have you found your supervision?

Probe:

Is it adequate and supportive?

Do you think this could be improved in anyway?

7) Have you been able to collaborate with other Community Navigators?

Probe:

If so, do you feel there have been any benefits of this?

8) Have you found there to be any challenges when doing this?
9) Were you included in any of the discussions regarding which clients you would be allocated during your involvement?

Probe:

Can you tell me how you felt about this process?

10) Would you have liked to have been involved in this more?
11) How have you found working with your clients so far?

Probe:

Is there anything you have found particularly difficult? i.e. connecting /communicating with clients

12) Have you noticed any barriers to people being able to connect with you? If so, can you tell me about these?
13) For any clients you have not met or have dropped out, what do you think contributed to this?

Probe:

Did you notice anything that indicated this was going to happen?

14) We know some people struggle to stay engaged with the programme, is there anything we could do differently to make the programme more engaging for clients?
15) Can you think of anyway that you think we could improve the programme?

### Internal Pilot Qualitative Interviews

### Supervisors

**This is a flexible guide to help build a conversation. Not all questions will need to be asked and may not be relevant to all interviewees. Interviewers will paraphrase question to best suit the person they are speaking with.**

**The purpose of this guide to is to *explore people’s experiences of the programme from the recruitment process, obstacles to initial participation, and any barriers to subsequent engagement.***

1) Can you tell me how you got involved in the study?

Probe:

How did you find this process?

Is there anything we can do to improve the initial involvement process?

2) Can you tell me about the training you received?

Probe:

How helpful did you find the training?

3) Is there anything we can do to try and improve the training / anything you think the training programme missed?
4) Do you feel your role and expected work of the Community Navigators was explained clearly enough?

Probe:

If not, what could be improved?

5) How do you think the recruitment of Community Navigator clients for the study is going?

Probe:

If struggling to recruit, why does the supervisor think this is?

Is it diverse and inclusive enough? If not how could it be improved?

6) Would you be able to tell me a bit about how you have been approaching supervision with your Community Navigator?

Probe:

How do you feel this is going?

How is communication between you and the Community Navigators? (Do you offer different approaches?)

7) Are there any barriers in providing supervision to the Community Navigators?

Probe:

If so: Could you tell me about these?

Can you tell me about anything we can do to reduce these barriers?

8) Would you be able to tell us a bit about the kind of issues the Community Navigators have been raising with you?
9) For any clients yet to meet their Community Navigator or have dropped out, what do you think may have contributed to this?
10) Is there anything that you think the programme may be able to do differently, to try and help those who have found it hard to engage with the programme so far?
11) Is there any other feedback you have so far which we haven’t covered?

### 2. Survey questions

### Qualitative Study One Survey

***The survey will be sent via an email invitation and will let people complete it online anonymously. It will be sent to people still engaged with the study and people who have disengaged to explore the experience of the recruitment process and any areas where information was unclear or off-putting and seeking recommendations for greater inclusivity in our messaging and approach.***

**Thank you for joining the Community Navigator programme in [name of site].**

1) Were you allocated to receive support from a Community Navigator by our study team [first and surnames names of the local CN team] Y/N

**If Yes:**

**We are going to ask you some questions about the way we communicated programme information with you.**

1.1) Can you tell us what interested you about taking part in the Community Navigator programme on first hearing about it? (FREE TEXT)
1.2) The amount of information the person who told me about the study gave me was:
- Too little
- The right amount
- Too much
  1.3) When you heard about the possibility of working with a Community Navigator for 6 months, supporting you to manage your mental health and experiences of loneliness, how did you feel about this offer: all that apply

Interested:	very	somewhat	slightly	not at all

Anxious:	very	somewhat	slightly	not at all

Worried:	very	somewhat	slightly	not at all

Optimistic	very	somewhat	slightly	not at all

Excited	very	somewhat	slightly	not at all

Other [free text]	very	somewhat	slightly	not at all

*If Anxious/Worried/Other selected: Can you tell us any ways that we could help with reducing these feelings about the study?*

1.4) The information flyer about the Community Navigator programme was clear and easy to understand [picture to be included]
- Strongly Agree
- Agree
- Neither Agree Nor Disagree
- Disagree
- Strongly Disagree
- Don’t know / can’t remember receiving this flyer
  1.5) The full information sheet documenting the research study and what was involved in taking part was clear and understandable
- Strongly Agree
- Agree
- Neither Agree Nor Disagree
- Disagree
- Strongly Disagree
- Don’t know / can’t remember receiving this information sheet flyer
  1.6) The researcher clearly explained the study and answered my questions.
- Strongly Agree
- Agree
- Neither Agree Nor Disagree
- Disagree
- Strongly Disagree
  1.7) Have you had any contact with your Community Navigator yet Y/N?

**If no:** *Can you tell us why you have not yet had any contact with your Community Navigator? (FREE TEXT)*

1.8) Have you met in person your Community Navigator yet Y/N?

**If yes:**

*How many times have you met with your Community Navigator?*

- 1-2
- 3-4
- 4-5
- 6 or more
  1.9) My Community Navigator has communicated with me: (multiple choice)
- On Zoom
- On telephone
- On text message
- Face to face
- Email
- Other [free text]
  1.10) I am happy with the method my Community Navigator has used communicated with me
- Strongly Agree
- Agree
- Neither Agree Nor Disagree
- Disagree
- Strongly Disagree
  1.11) The amount of communication between me and my Community Navigator so far
- Too little
- The right amount
- Too much
  1.12) I have had any questions I have had answered so far by my Community Navigator
- Strongly Agree
- Agree
- Neither Agree Nor Disagree
- Disagree
- Strongly Disagree
- Not Applicable
  1.13) My expectations of the Community Navigator programme so far have been met.
- Strongly Agree
- Agree
- Neither Agree Nor Disagree
- Disagree
- Strongly Disagree
  1.14) Do you plan to continue working with your Community Navigator until the end of the programme Y/N
  1.15) Did you have any concerns about starting the Community Navigator programme? If so, can you tell us about these? (FREE TEXT)
  1.16) Have you encountered anything that has stopped you from engaging with your Community Navigator? If so, can you tell us about these? (FREE TEXT)
  1.17) Any other feedback on how we have set the programme up, and your experiences to date that will help us improve this project – please provide below (FREE TEXT)

**If No:**

**People who are part of our study but not seeing a Community Navigator are vital for this research. Your answers will help us improve how we communicate with you over the next 2 years.**

2.1) The information flyer about the Community Navigator programme was clear and understandable [picture to be included]

- Strongly Agree
- Agree
- Neither Agree Nor Disagree
- Disagree
- Strongly Disagree
- Don’t know / can’t remember receiving this flyer

2.2) The full information sheet documenting the research study and what was involved in take part was clear and understandable

- Strongly Agree
- Agree
- Neither Agree Nor Disagree
- Disagree
- Strongly Disagree
- Don’t know / can’t remember receiving this flyer

2.3) The researcher clearly explained the study and answered my questions.

- Strongly Agree
- Agree
- Neither Agree Nor Disagree
- Disagree
- Strongly Disagree

2.4) Did you have any concerns about taking part the Community Navigator study? Can you tell us about these? (FREE TEXT)

2.5) How did you feel not being allocated to meet with a Community Navigator?

Relieved:	very	somewhat	slightly	not at all

Anxious:	very	somewhat	slightly	not at all

Disappointed:	very	somewhat	slightly	not at all

No strong feelings	very	somewhat	slightly	not at all

Other [free text]	very	somewhat	slightly	not at all

2.6) How well did the research team staff communicate the information that you would not see a Community Navigator to you?

- Poor
- Fair
- Good
- Very good
- Excellent

*How could we have improved this?*

2.7) Have you accessed or found out information about other community supports or local resources? Y/N

*Could you tell us about these?* (FREE TEXT)

2.8) Any other feedback on how we have set the programme up, and your experiences to date that will help us improve this project – please provide below (FREE TEXT)

**3. Survey responses to closed questions**

| Question | Total responses | Too much | The right amount | Too little |
| --- | --- | --- | --- | --- |
| 1. The amount of information I was told about the study was | N=14 | 1 | 12 | 1 |
| 2. The amount of communication between me and my Community Navigator so far is | N=13 | 0 | 13 | 0 |

| Question | Total responses | Strongly agree | Agree | Neither agree nor disagree | Disagree | Strongly disagree | N/A |
| --- | --- | --- | --- | --- | --- | --- | --- |
| 3. The information flyer about the Community Navigator programme was clear and easy to understand | N=23 | 10 | 8 | 5 | 0 | 0 | 0 |
| 4. The full information sheet documenting the research study and what was involved in taking part was clear and understandable | N=23 | 7 | 12 | 3 | 1 | 0 | 0 |
| 5. The researcher clearly explained the study and answered my questions. | N=23 | 16 | 7 | 0 | 0 | 0 | 0 |
| 6. I am happy with the method my Community Navigator has used to communicate with me | N=13 | 8 | 5 | 0 | 0 | 0 | 0 |
| 7. I have had any questions I have had answered so far by my Community Navigator | N=13 | 6 | 5 | 1 | 0 | 0 | 1 |
| 8. My expectations of the Community Navigator programme so far have been met | N=13 | 3 | 5 | 4 | 0 | 0 | 1 |

| Question | Total responses | Relieved | Anxious | Disappointed |
| --- | --- | --- | --- | --- |
| 9. How did you feel not being allocated to meet with a Community Navigator? | N=9 | Not at all = 8  Slightly = 1 | Not at all = 6  Somewhat=1  Very = 2 | Not at all = 2  Slightly = 2  Somewhat = 1  Very = 4 |

| Question | Total responses | Excellent | Very good | Good | Fair | Poor |
| --- | --- | --- | --- | --- | --- | --- |
| 10. How well did the research team staff communicate the information that you would not see a Community Navigator to you? | N=9 | 3 | 2 | 2 | 1 | 1 |

**4. Thematic framework**

| **Community Navigator Themes & Subthemes** | | |
| --- | --- | --- |
| Theme Name | Sub themes | Quotes |
| Training & Preparation | Positive experiences   - Involvement of LEAP / role play scenarios - Access to manual & training videos   Negative experiences   - Large gap between training and seeing SU’s   NHS culture:   - Integration & navigation issues - Lone working concerns - Tech issues (= Barrier to engagement with Sus) | *I thought what was particularly helpful was some of the scenarios that we did as well.*  *And also some of the training we did where there were some lived experience consultants there as well and their input was really valuable and to hear some of their ideas as well*. (CN7)  *There was about a six-month gap for me between the training and meeting people. What was very helpful about the training was that we were sent manuals and resources so that I could tap back into that and go back over it before I went in to support people. So yes, I did feel like it prepared me well*. (CN6)  *I suppose it’s just a shame that we weren’t actually seeing anybody and had anything to go on, in that sense, when you were thinking about some of the scenarios and things. But, for me it was a positive experience.* (FG3)  *I think that was just how everything is set out to be. It was a little bit scattered in that sense, but I think this probably was my personal experience, maybe because of all the issues that I have had with starting the actual job, (Laughter) because it felt quite a long time before I actually started*. (CN8)  *…then I’ve got a week where I've been ordered to get onto the NHS training, I've got a two-day face-to-face training course. So it's going to be quite a gap before I see her again.* (FG1)  *"but I think I definitely initially struggled with the system, with the NHS, because it was all different. I didn't have prior experience of working for the NHS, so just, sort of, to get all the training out of the way and just... I think the first few times I went into the office was overwhelming because everyone seemed to be knowing what they were doing and where they go, and I just felt a bit lost, I think, for the first few weeks."* (CN2/ FG1)  *"There's so much essential training within the NHS. I was a bit overwhelmed by how much online training and face-to-face training there is to do for a one-day-a-week course."* (CN1 / FG1)  *But I have worked in a team before in a trust where, like, in the team, it was, like, when we were working from home and things, we just got a call in the morning and a call at the end of the day...Whereas I haven't felt like I've really known, if I'm supposed to be doing that with anyone, and if so, who's it with? Kind of thing*. (CN7)  *But I think definitely it did make me conscious of lone working policies and how this is definitely not ideal, safety-wise, or not being in contact with my line manager if I need something.* (CN2 /FG1)  *"So, I had some snags with my work phone for a long time, I got my work phone this month, literally....I think one of the things if I could just say, was just me not having a work phone for a while. That was a challenge because sometimes they couldn't really get to me or if I called them, they couldn't call me back because I would call from a private number. So they couldn't get back to me and confirm appointments or, sort of, have that contact with me, or receive text message reminders."* (CN2 / FG1) |
| Supervision & Support | Positive experiences   - Frequent meetings - Working the same days as CN - Supervisor qualities   Negative experiences   - lack of initial support - insufficient knowledge of CN role - absence of supervisor - multi-disciplinary team meeting (MDT) | *Yeah, mine’s been regular. It’s been every fortnight, and that’s one of the really positive relationships that I’ve got outside of the community navigator group, is my supervisor and the relationship that we’ve got*. (FG3)  *Mine's been a really good experience. I work generally the same day as the person that’s supervising me...But yeah, I've had quite a lot of supervision and it's been really helpful in terms of at the beginning it made a massive difference for me in terms of navigating myself, the NHS, really, and understanding the systems because it is very different to other systems.* (FG1)  *I think (supervisor) is nice, but he is not too directive. He is quite gentle as a supervisor, I guess...* (CN7)  *I feel like she’s approachable. I can share things with her...(*FG3)  *My induction there was I met with (supervisor). We had a short meeting because he was a bit late, and then he had to leave so I joined the team meeting with everyone at (site name) which was helpful...After that, he wasn't there to discuss it, let's say, or that was kind of it. (Laughter) That was kind of- Then the rest, I found out myself what I had to do in terms of training.* (CN8)  *I don't know, I get a sense that he is finding his feet, too, with this programme, but maybe it is just my perception.* (CN8)  *I was kind of just left to my own devices without any help, and this is why I felt so isolated. I had to ask for another supervisor, because I could see she was clearly going to be off for a long time. So, I had no one to go to, really. Yeah, so I had to ask (NHS Trust Team Member) for somebody, which eventually they did give me somebody, but it was only like once a month, and quite brief, really. And I just got the impression that they’re just so inundated and got so much work that you were kind of just left to it, to get on with it.* (FG3)  *"But then she's also really, really busy. I know that because she manages the entire West locality and probably South locality teams. But yeah, in terms of if I've had other questions, I couldn't really say that I could go to anyone, I suppose."* (CN2 / FG1)  *It's been quite helpful being part of MDTs because that way you can, kind of, keep refreshing people about the project and letting them know it's running...And when new people join, you know, they're aware as well because you get that chance to talk about it. So that's been helpful as well. And I think that's probably quite an important part of the project.* (CN7) |
| Team Collaboration | Team building  Support  Challenges   - staggered starting times - working different days / different locations | *And I think me, (Community Navigator 1) and (Community Navigator 4) I feel like we’re a team. And we’ve got a little WhatsApp group, and we message on that, and we support each other. So, from that respect, that’s really positive, that we all get on really well and we’re there for each other.* (FG3)  *"We've got a WhatsApp group, the three of us, on a, kind of, private phone so that we can communicate, not about clients, but about any arrangements. And we've gone out and done visits together to (Charity Name), which is a women's service where we are, and (Mental Health Charity Name), which is a mental health education service as well. So, we've gone and done that together, which has been really nice."* (CN1 / FG1)  *"So, I don't have any colleagues in (Site Name) yet. Hopefully soon."* (CN2/FG1)  *"I'm in the same office as one person but working on a different day and I know they work the same day but in different offices*." (CN1/FG1)  "*for example, this week, because I'm collaborating with my other two community navigators, and seeing my person on Friday, it means I'm working three days."* (CN1/FG1) |
| Key Traits / Skills | Trustworthy  Honesty  Rapport Building  A good listener  Reassuring & confidence building  Empathetic, understanding, compassionate | *…in the notes she'd spoken to another member of staff where it was written in the notes that it was going well with the community navigator and, you know, it had been suggested about meeting for a coffee rather than her home address.*  *And she was keen to try it because she felt she could trust the community navigator. And so that's really nice feedback, but at the same time, it, kind of, underlines that importance of trust*. (CN7)  *And one thing I said to every client in the first session is, “Look, I'm a human being and I'm going to say things sometimes which aren't helpful.” And I always say, “Look, I want this to be based on honesty. So if I say something that's not helpful or do something that's not helpful, please tell me because that's the only way I learn about you and what works for you best.” And I think that approach has been really helpful in, kind of, building up that trust and them knowing, you know, they can say to me, “Actually, I don't like that and that is really not helping me.” And I have had clients who've said that to me. So that's been quite positive really because... they've never said it in a bad way. It's more like we've explored it together a bit.* (CN7)  *"I've realised, actually, if you are going to be the connector, you do actually need to have a connection with the person. There’s a lot of trust involved, which I don't think I appreciated initially. And I think it's a very fine line and you have to be careful between being, you know, the actual role that you're doing and then not doing too much for someone."* (CN7)  *I find that some clients like to use the space that we have to talk quite a lot about what's going on in their lives generally, and things that they're not happy about. So I always allow a bit of space for that to happen. Even though I know that technically that's not the role, but I think actually me being- listening to how they're feeling and what's going on for them in their lives and the things they're worried about, kind of, builds up some of that trust, which I think is really helpful when we, kind of, get into a stage where I am now with clients where we're going to try going to things.* (CN7)  *She was quite concerned that she would somehow fail because she felt that she might not do as much as this programme requires or what the idea is to make them. .. So it was, like, a lot of reassuring her that, “There's no success or failure and that we will be able to plan things how you want, and goals can change.” And, you know, “There's really no hard and fast, “You have to succeed at this”.” But I think it was reassuring to her and she was quite happy to, like, meet up.*  *"My client has said that he only went out to put the bin out. I mean, he has gone out. He’s actually gone to a football match and done a few other bits and pieces, but that’s with friends he’s known for a long time. But, from the point of view of actually getting out there and going to meet people he doesn’t know, it’s a massive step for people, isn’t it, often?.., It kind of proves that they can do it, I suppose, as well, doesn’t it? And then it’s not as bad as they think it might be, especially when you bump into a friend that you can have a good chat with."* (FG3) |
| Challenges faced with working with service user’s. | Complex issues  Disengagement  Boundaries (in terms of):  - Flexibility  - Emotional  Staying on track | *just because I am working with quite complex people at the moment, I have been wondering about boundaries...here the boundaries are quite blurred...I guess the challenging part for me also was not to go into therapy as well, like to be empathic, but also, again, just reflecting about my role and keeping it safe for everyone, really*. (CN8)  *But every time, she would cancel it due to physical health problems or physical health appointments. When she did withdraw from the project, I did wonder if there was anything I could have done more at that point to, kind of, try to get her to meet in person. And there was part of me that wondered, if we had been able to meet in person, if that might have helped her to stay in the trial.* (CN7)  *"So, I'm happy to do it and I have done it a bit, to where I can, but there are limits because of other commitments really. That's probably the biggest challenge is how you maintain really positive contact but don't let it take over, in a sense, and have a few boundaries in place really."* (CN1/FG1)  *just because I am working with quite complex people at the moment, I have been wondering about boundaries...here the boundaries are quite blurred...I guess the challenging part for me also was not to go into therapy as well, like to be empathic, but also, again, just reflecting about my role and keeping it safe for everyone, really.* (CN8)  *"I try to bring the focus back to what we are doing, but sometimes it may not be entirely appropriate, but just somehow, you know, to keep things going at least. But I do worry that it might mean if someone’s struggling a little more in the future, it might be hard to just define goals and work on them and just, sort of, get that feedback."* (CN2/FG1)  *"But then they also have bad weeks sometimes, and if we have had a session during that time, it's just been hard to plan anything because you obviously can't when the person's really distressed and probably just wants to talk."* (CN2 / FG1) |

| **Service User Themes and Subthemes** | | |
| --- | --- | --- |
| **Theme Name** | **Subtheme** | **Quotes** |
| Personalisation & Choice | SU's having preference over their communication methods with CN.  SU’s given a choice of which format information is provided. | "And the facility that I can text her, or for example if I miss a call I can text her or she can reply to me, she can phone me back, something like that. Simple things like that, so it’s been okay." (SU4)  "Yes, the information was presented verbally on the phone, and then on paper form. I had everything I needed to make an informed choice. No, I would say, yes, wouldn’t really say there is anything else you could do differently or better." (SU10) |
| Flexibility | Essential for CN to be flexible regarding service user's preferences.  Limited CN working hours - not always available and able to be flexible when SU needs.  Ability to work with individuals’ accessibility needs. | "And she always goes that extra mile, if I don't want to meet over there, she'll say, we'll go for coffee, or we'll go for a walk or something like that. So she's always there, you know?" (SU4)  "...because she works part -time, obviously, she’s not contactable, what is it? Thursdays and Fridays, and whilst I’ve been okay to meet, I have to reschedule. And, yes. I mean, it is a bit of a pain having to make a mental note set a reminder to do it Monday. But I suppose you can’t really do much about that." (SU11)  "Yes, she's really good. She's really helpful. So, she's really accommodating. She has said, "We'll do as very little as you want." Fully understands what's going on. If I need to video call with her. If I've had a crash or something like that, she's absolutely fine with it. Yes, just very, very understanding. I'll be completely honest. She's been more understanding than some therapists that I've come across while accessing certain services. So, she's been really, really helpful." (SU13) |
| External Barriers | Trustworthy initial introductions: essential for CN programme to be introduced correctly by their care coordinator so they know what they are singing up for. | "but my expectation was that maybe they’re like a support worker, maybe she is like a support worker. She is dealing with my needs every day...And (NHS trust service) didn’t care, didn’t listen to me, and didn’t help me. She kept saying, “Now, you have a care coordinator. Ask her.” And that’s why it made me more mad, it made me more sad. And I just came out of the research, you know, programme. And I said, “I want to withdraw”, because it’s going to be more complicated for me." (SU16) |
| Internal Barriers | Self-beliefs - SU's questioning if they are deserving of the programme.  Current MH difficulties can make engaging with CN and trying new things hard.  Timing - for the programme to be useful, the SU needs to be at a stage of wanting to find new social connections. | "I think my only thing was, I guess in some ways it's probably just my own brain but, “Am I too well for this study”, kind of stuff" (SU5)  "because my mental health isn't there basically, just isn't there on point, and then I end up not engaging as much as I probably should, or I wasn’t able to respond and stuff like that." (SU15)  "But sometimes when you have a depression, high level of anxiety and stress, you can’t focus on what you’re hearing, what you’re learning. Sometimes I need to ask the people to repeat the words many times." (SU16)  "But, you know, it’s very hard, when you’re depressed, or you have a mental health problem, to go and find your hobbies, or anything. And that’s why it was really hard for me to continue with the care navigator. Just thinking about it, what I am going- because at that moment, I didn’t enjoy anything, you know? ...And there is no point for me to talk about socialising" (SU16) |
| Kindness & Patience | Treated with kindness & patience by first member of staff, believe future engagement with other staff members will be the same = positive expectations  CNs described as understanding - SU's often seemed surprised by this - suggest a novel & unique approach.  kindness of CNs allows SUs to feel comfortable around them.  SU being in control of the pace ensures comfort: person centred approach.  Research assistants treated service users with only kindness and patience. They were understanding and clear in their explanations. Always open to questions. | "I think the Community Navigator that I'm with was very understanding. It was, so you feel like you are being listened to really, so that is good...Yes. I thought it was probably going to be one of those on treatment team visit where they just ask you routine questioning and that is about it, and stuff like that." (SU15)  "I find it’s okay, I find easy things, because to be honest, I have an amazing person with me, my navigator....I’ve been able to speak and been able to feel comfortable and to have the things I expect to happen, to have the amazing person with me, to not feeling like, how can I explain? I’m feeling so comfortable with that... It’s been amazing. I mean she’s been so helpful because I said when I have somebody to express myself and to speak of this, someone that can understand, I feel so good, I feel so comfortable" (SU3)  "She said that she's there for me. She doesn't want to push anything on me. It's more about just guiding me, rather than pushing anything on me. So, yes, she's just- yes, just very understanding..."(SU13)  "That experience for me was good, it was very good because I was always treated with patience, with kindness. Yes, that was important to me after being so hesitant because of the reasons that I said before. I took the decision, I get the first step, because she explained me clearly. She was very kind. She was patient. So for me it was a good experience" (SU3) |
| Contagion | (experiences at one stage of the recruitment process affect attitude towards the next stage)  Positive  Negative  Some SU’s had negative feelings towards being allocated into the control group. This may affect their later involvement in the study. | "I usually normally feel nervous, but I have to be honest, I was expecting to meet a very kind person, understanding. Because when they told me it will be another person, and I said hmm, I remember thinking hopefully I will meet a nice person. So it was basically, to be honest, my expectation I really hope that I can meet them, my Community Navigator will be understanding and, yes, a nice person. And when I met her, (Community Navigator 3) it was, yes, it’s been okay, she’s a really nice person." (SU3)  "There was a lot of communication at first. It was quite, if I'm honest, bombarding. And I hadn't seen anybody at that point, I was like, “Wow.” I found it a bit overwhelming. Because I didn't really know... When I was getting people calling me, I wasn't aware, at the time, these were the people... I didn't know I was going to get called up all the time like that, so I found it quite overwhelming, because I really don't like the telephone" (SU8)  “Disappointment after rejection time after time have negatively impacted my mental health leading to self-harm AGAIN and more time in A&E” (Survey Data).  When asked “How did you feel not being allocated to meet with a Community Navigator?” 4/9 participants reported to be very disappointed, 1/9 somewhat disappointed & 2/9 slightly disappointed. 1 participant also reported “Feelings of loss and that I was not good enough”. (Survey Data) |
| Fine Balance of Information | This refers to the amount of information the SU is given during the recruitment process.  Just the right amount.  Missing Information.  Information overload.  a lot of information to take in - this can be hard when struggling with concentration.  Essential to ensure someone is there to help the SU. | "No, I feel like I had enough information,...but sometimes I can get overwhelmed with too much information about it, I was given enough information that I understood what it was, but didn't need to know every single intricate detail. But also knew that I could ask questions if I needed to." (SU5)  "I know that my Community Navigator told me that that there's a pocket of funds to help you access things if you don't have the financial means to. I think that could have been quite helpful to know at the beginning." (SU5)  "Yes, it was easy to understand, but it is just my concentration isn't very good so it took me a little while, that is all." (SU15)  "A bit overpowering, because in your head, sometimes it gets a bit foggy, you know?...just make sure that there's someone there that can explain it properly and help fill the paperwork in, that's the only thing. I think a lot of people struggle with..." (SU4) |

| **Recommendations** | **Description** |
| --- | --- |
| Documented Personal Communication Preferences | Based on feedback from the survey about the experience of being recruited to the trial, we double-checked with participants at all stages about their preferred communication method (e.g., text, email, or calls), documented these preferences, along with their preferred name, for future reference. |
| Proactive Contact | Researchers told participants as soon as possible if plans changed or they were running late, because this was identified as a source of anxiety. |
| Sensitive Communication around Allocations | Allocations (TAU or CN) were communicated over the phone, with a pre-arranged time set by text. Participants are informed in advance that this call would reveal their allocation (so they were not blindsided and could arrange support). We explained the process and emphasised that the allocation is not a reflection of their current mental health and that it would not affect they care they received from teams: that it was completely random. |
| Clear Expectations | The research team carefully described the support available to ensure participants understood the intervention without over-promising on flexibility. Researchers were encouraged to check information (e.g. about Community Navigators’ availability and work hours) rather than commit to uncertain details. |

**5. Feedback adjustments**
